# White matter brain age as a biomarker of cerebrovascular burden in the ageing brain

**DOI:** 10.1101/2022.02.06.22270484

**Authors:** Jing Du, Yuangang Pan, Jiyang Jiang, Ben C. P. Lam, Anbupalam Thalamuthu, Rory Chen, Ivor W. Tsang, Perminder S. Sachdev, Wei Wen

## Abstract

As the brain ages, it almost invariably accumulates vascular pathology, which differentially affects the white matter. The microstructure of the white matter may therefore reveal a brain age reflecting cerebrovascular disease burden and a relationship to vascular risk factors. In this study, a white matter specific brain age was developed from diffusion weighted imaging (DWI) using a three-dimensional convolutional neural network (3D-CNN) deep learning model in both cross-sectional data from UK biobank participants (n = 37327) and a longitudinal subset (n = 1409) with an average of 2.25 years follow up. We achieved a mean absolute error (MAE) of white matter brain age prediction of 2.84 years and a Pearson’s r of 0.902 with chronological age in the test participants. The average white matter brain age gap (WMBAG) of the baseline 1409 participants with repeated scans were 0.36 ± 0.11 years younger than that of other participants in the baseline test sample with single time-point MRI scan (n = 9759). Individual vascular risk factors and the cumulative vascular risk score were significantly correlated with greater WMBAG Obesity was observed to be correlated with WMBAG only in the male participants. Participants with one, two, and three or more vascular risk factors, compared to those without any, showed an elevated WMBAG of 0.54, 1.23, and 1.94 years, respectively. Baseline WMBAG was also associated significantly with processing speed, executive and global cognition after Bonferroni correction. The significant associations of diabetes and hypertension with poor processing speed and executive function were found to be mediated through the WMBAG. However, the vascular risk factors did not associate with the two-year change in WMBAG in the longitudinal dataset. Our analysis suggests that tissue-specific brain age can be successfully targeted for the examination of the most relevant risk factors and cognition, although longer-term longitudinal data are needed to demonstrate its dynamic characteristics. The results suggest an intriguing possibility that a white matter brain age gap can serve as a potential neuroimaging biomarker for an individual’s cerebrovascular ageing process.

## 1. INTRODUCTION

Brain ageing is a complex biological process in middle-to-late aged individuals, accompanied by changes at all levels from molecules and morphology to advanced brain functions^1^. Exposure to different hazardous vascular risk factors such as hypertension, diabetes, and hypercholesterolemia aggravate the vascular burden and accelerate the cerebrovascular ageing trajectory. Given that different organs or systems usually demonstrate heterogeneous ageing rates, an individual might have multiple underlying bodily ages^2^, such as bone age, renal age, and lung age, in addition to their chronological age. Brain age is a special case in this context, and it arguably reflects the brain’s biological status. Previous studies have investigated brain age using high-dimensional neuroimaging data in a machine learning framework for healthy populations ^3, 4^ or people with specific brain diseases ^5, 6^. Brain age gap (BAG) is the difference calculated by subtracting the chronological age from predicted brain age, which describes how one’s brain health deviates from what would be expected for the chronological age. Positive brain age gap (i.e., an older predicted brain age than chronological age) was usually reported to be associated with brain disorders such as Alzheimer’s Disease (AD) ^7^, multiple sclerosis ^8^, and schizophrenia ^6^.

Different anatomical locations and brain tissue types of the brain show different vulnerability to vascular risk factors^9, 10^. Due to the ‘outside in’ vascularisation pattern^11^ with key regulating vessels outside the brain parenchyma, the number of arterioles supplying grey matter are considered eight times more than that in white matter^12^. As a result, white matter, particularly deep white matter, is more susceptible to ischaemia than grey matter. Brain lesions observed in cerebrovascular disorders are more relevant to white matter, such as white matter hyperintensity (WMH), lacunes, microbleeds and enlarged perivascular spaces^13^. Therefore, to investigate the cerebrovascular burden on the white matter, tissue-specific brain ages should be established. To our knowledge, most brain age studies thus far have generated a single brain age using T1-weighted imaging scans. Tissue-specific brain ages, especially white matter brain age, have not been fully investigated. A recent study ^14^ proposed a grey matter brain age, which was suggested as a biomarker of the risk of dementia. Benson Mwangi et al.^15^ developed a diffusion weighted imaging (DWI) derived brain age, but the main aim of the study was to prove the concept and demonstrate the validity of establishing brain age using DWI scans rather than the usual T1-weighted scans. Further, they did not explore specific relationships between cerebrovascular burden and this DWI-derived brain age. In a more recent study^16^, over 1000 neuroimaging phenotypes derived from multimodality MRI scans including DWI were used in a linear regression model to estimate brain age. So far, there has been few explorations of the relationship between vascular risks and its cerebrovascular consequences in white matter by using white matter brain age.

The primary objective of this study was to investigate whether vascular risk factors would be associated with an accelerated brain ageing process measured by ‘white matter specific brain age’ in community-dwelling, middle to older aged adults drawn from a UK Biobank cohort cross-sectionally and a longitudinal subset. DWI is a sensitive and reliable technique for monitoring white matter microstructural impairment. Using deep learning techniques, we developed a DWI-derived white matter brain age as a biomarker to characterise the microstructural changes in relation to vascular risk factors and cognition. The individual and accumulative effects of vascular risk factors and their sex stratifications on the white matter brain age gap (WMBAG) and cognition were examined. We hypothesised that the DWI-derived white matter brain age would reflect the cumulative cerebrovascular burden and thereby enhance the understanding of how vascular risk factors may accelerate the biological ageing of the white matter.

## 2. METHODS

### 2.1 Participants

Data for this study were drawn from UK Biobank, a large-scale ongoing prospective population-based cohort study ^17^. A flowchart of the selection of participants can be found in Figure 1. Briefly, after visual inspection of 37327 eligible DWI scans, 98 participants with poor image quality were removed, leaving 37229 at baseline to be included in this study. After excluding 3399 participants with severe self-reported brain related disorders (Field ID 20002, see Supplementary Table e-1) to ensure a relatively healthy sample for deep learning training, 60% (n = 19526) of the remaining participants were randomly selected to the training set. Twenty percent (n = 6515) were used as the validation set, which provided an unbiased evaluation of a model fit on the training dataset while selecting the model’s structures (e.g., the type of loss function and optimiser for training a neural network). The remaining 20% were combined with the unhealthy participants as identified above (n = 11168) for use in the test set. In this test sample, 1409 participants had both baseline and follow-up scans that were used for longitudinal analysis.

**Figure 1.**
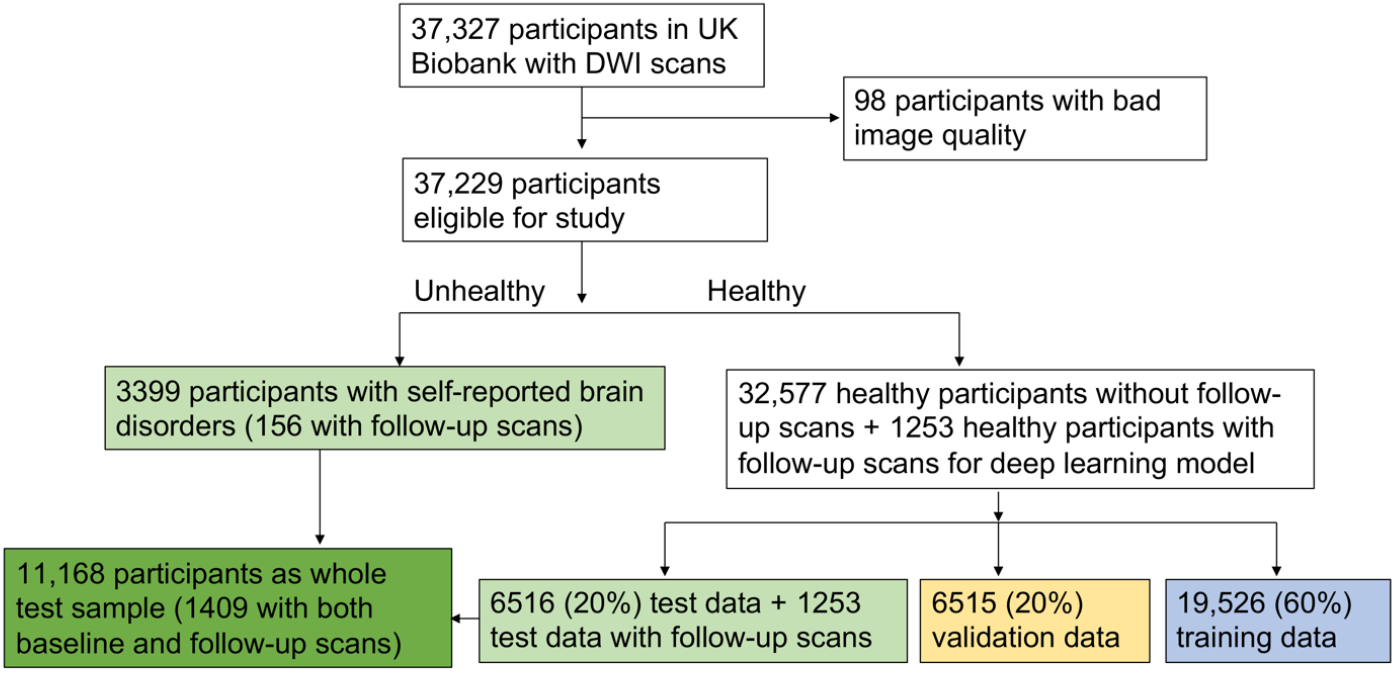
Flowchart of participant selection.

The ethics of this study has been approved by the North West Multi-centre Research Ethics Committee (MREC) and written informed consent was obtained from all participants.

### 2.2 MRI acquisition and imaging processing

DWI scans were acquired from three imaging centres (Cheadle Greater Manchester, Newcastle and Reading, UK), and each centre used a 3T Siemens Skyra scanner with a standard Siemens 32-channel head coil and same parameters. Details of imaging protocols can be found in the online UK Biobank brain imaging documentation (https://biobank.ctsu.ox.ac.uk/crystal/crystal/docs/brain_mri.pdf). The original DWI data had been pre-processed with eddy currents and head motion correction by UK Biobank using the FMRIB Software Library (FSL) toolkit^18^. The diffusion-tensor-imaging fitting tool (DTIFIT) was used to generate the following DWI-derived maps in native space: FA (fractional anisotropy), MD (mean diffusivity), AxD (axial diffusivity), RD (radial diffusivity) and MO (tensor mode). The individual maps were nonlinearly warped to a 2 × 2 × 2 mm^3^ MNI-152 standard space using FNIRT (FMRIB’s Nonlinear Image Registration Tool)^19^. The standard-space DWI-derived maps for each individual were finally used as the input for the deep learning model. All DWI-derived maps were visually inspected before being used as input for the deep learning networks.

### 2.3 White matter brain age computation

A three-dimensional convolutional neural network (3D-CNN) deep learning model was used to establish white matter brain age, the structure of which is illustrated in Figure 2.

**Figure 2.**
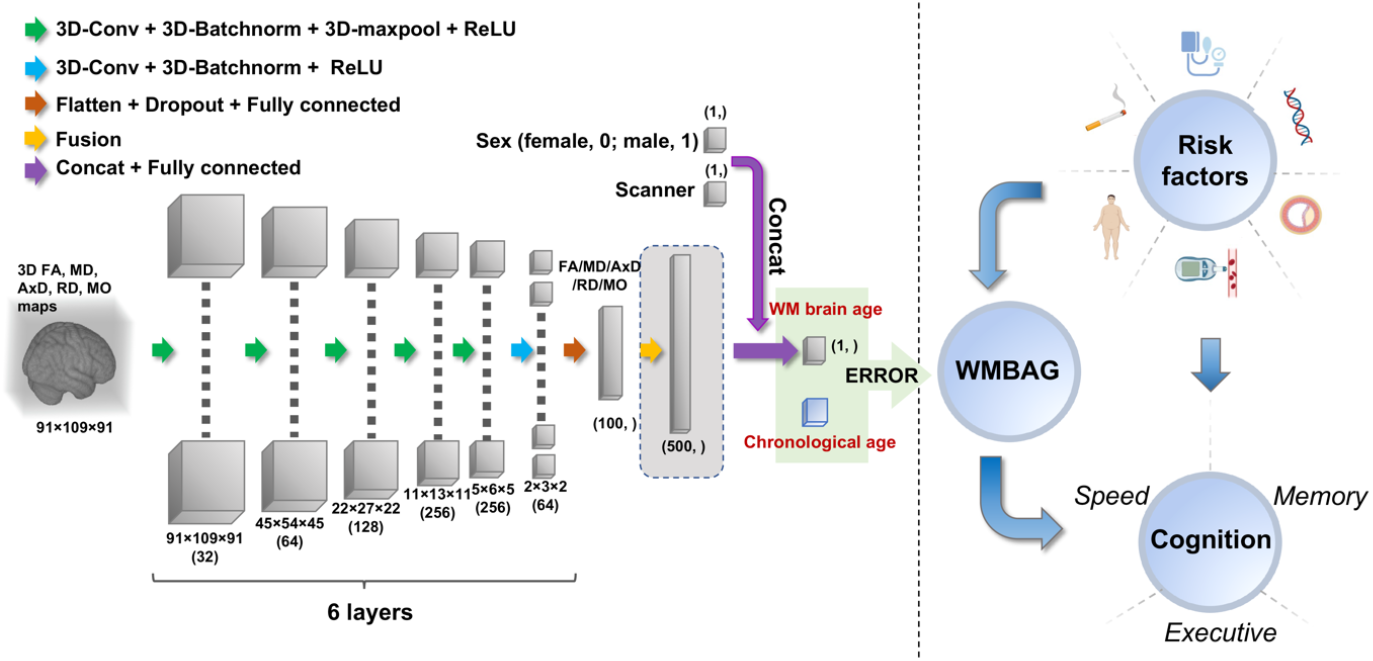
Overview of study design. The left panel shows the 3D convolution neural network (3D-CNN) architecture; the right panel shows the clinical analyses between risk factors and WMBAG and cognition. Inputs of the model are pre-processed 3D FA/MD/AxD/RD/MO maps, WMBAG = White matter brain age – Chronological age. Abbreviations: 3D = three-dimensional; Conv = convolution; Batchnorm = batch normalization; ReLU = rectified linear unit; WM = white matter; WMBAG = white matter brain age gap; FA = fractional anisotropy; MD = mean diffusivity; AxD = axial diffusivity; RD = radial diffusivity; MO = anisotropy mode.

#### 2.3.1 Convolutional neural network architecture for each DWI map

A 3D-CNN architecture was applied to estimate white matter brain age using DWI maps. The architecture follows the Simple Fully Convolutional Network (SFCN) proposed by Peng Han et al. ^20^ due to its simplicity, which is based on VGGNet ^21^ with fully convolutional structures^22^.

Briefly, the network received a 91 × 109 × 91 3D image and the corresponding sex and scanner of a participant as input, and the output was the predicted age at the last layer. The network consisted of eight blocks, as shown in Figure 2, and each of the first five blocks contained a 3D convolutional layer with kernel size 3 × 3 × 3, a 3D batch normalisation layer, a 3D max-pooling layer, and a ReLU^23^ activation layer. The sixth block had a 1 × 1 × 1 3D convolutional layer, a batch normalisation layer, and a ReLU activation layer. The seventh block contained a dropout layer (activated only during the training process by randomly dropping 50% of the elements) and a fully connected layer. The spatial dimension was reduced to 2 × 3 × 2 after the sixth block. The flatten operator was applied to resize the tensor to a vector before applying the seventh block. Instead of going through the 3D-CNN network as images, the extra information such as sex and scanner was incorporated into the feature map by concatenation before the eighth block. Finally, linear regression was employed in the eighth block for fusing the image features and the information of sex and scanner, with the output being a scalar for the predicted white matter brain age. The channel numbers used in the first six 3D convolution layers were [32, 64, 128, 256, 256, 64].

The internal process of the model can be summarised into three stages: (1) *Nonlinear feature extraction*: The first six blocks extracted feature maps from each input image; (2) *Tensor to vector*: The seventh block smoothly transformed the 3D tensor to a vector for downstream age prediction; and (3) *Linear regression*: The eighth block incorporated the extra sex and scanner information, and the output was the predicted age.

#### 2.3.2 Network architecture for fusing all five diffusion maps

To increase the white matter brain age prediction accuracy, the five resultant DWI-derived maps (FA, MD, AxD, RD and MO) were incorporated to generate a composite metric. Five 3D CNN networks with the same structure as discussed above were applied, with each network modelling each feature map separately. In terms of feature fusion, we adopted a simple concatenation to fuse the five feature maps after the seventh block, as well as the covariates (i.e., sex and scanner). The two covariates were applied to all five feature maps. The resultant feature map was a vector with (100*5 +2) entries, namely 100 entries for each feature map and 2 entries for two covariates. Similarly, linear regression was employed in the eighth block for mapping the fused features into the final predicted age.

To reduce the computational cost, instead of retraining five 3D CNN networks simultaneously from scratch, we reused the intermediate feature maps learned during the analysis of each of the five DWI-derived maps and only trained the eighth block accordingly.

#### 2.3.3 Bias correction

The predicted ages normally suffer from the issue of underfitting due to regression dilution and non-Gaussian age distribution, which means older participants will be estimated with a younger brain age while younger participants will be estimated with an older brain age. As reported by Smith et al.^24^, bias correction is an essential postprocessing technique in most brain-age prediction studies. Using the techniques proposed by Smith et al^24^, we applied the linear bias correction method to the predicated age before the subsequent investigations with clinical measurements, where *y* and *ŷ* denote the chronological age and predicted age, respectively. We can fit a linear regression *ŷ* = *αy* + *β* on the left-out validation set with known chronological age. Applying the learned coefficients (*α,β*), the corrected predicted age *ŷ*_*c0*_ for test set can be estimated by

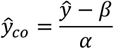

where we assume the coefficients (*α,β*) can be generalised to the test set.

#### 2.3.4 Model performance

Model performance was evaluated by two predominant measures in this study. Mean absolute error (MAE) was defined as 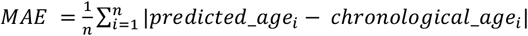, Pearson’s correlation coefficient (Pearson’s r) was applied to characterise the correlation between chronological age and predicted age.

### 2.4 Vascular risk score (VRS) and Apolipoprotein E (APOE) ε4 carrier status

While a variety of vascular risk factors have been reported in previous decades, we incorporated five essential vascular risk factors, i.e.: (1) hypertension; (2) diabetes; (3) hypercholesterolemia; (4) obesity; and (5) smoking. An Omron HEM-7015IT device was used to automatically evaluate the seated blood pressure twice; the mean blood pressure for each individual was computed by averaging these two measurements. Participants with hypertension were defined with blood pressure over 140/90 mmHg or using lowering blood pressure medication (Field IDs 6177 and 6153). Participants with diabetes were defined according to the doctor’s diagnosis (Field ID 2443) and anti-diabetes medication (Field IDs 6177 and 6153). Hypercholesterolemia was identified according to the medication information (Field IDs 6177 and 6153). Obesity was defined as body-mass-index (BMI) ≥ 30, which was constructed from ratio of height and weight measured during the initial assessment (Field ID 21001). Smoking was defined as current or previous smoking history (Field ID 20116). Each vascular risk factor was binarized with 1 indicating presence of that factor and 0 otherwise. A composite vascular risk factor score (VRS) was generated to evaluate the overall cerebrovascular burden by calculating the total numbers of vascular risk factors using a method applied similarly in other studies^25^. Given that there were a very small number of subjects who scored 4 (n = 408, 3.7%) or 5 (n = 82, 0.7%), the VRS was categorised into 0,1, 2 and ≥ 3.

DNA from a blood sample of first recruited participants (approximately 50,000) were genotyped in the UK biobank using Affymetrix UK BiLEVE Axiom array; the rest were genotyped with Affymetrix UK Biobank Axiom array^26^. Two APOE coding single nucleotide polymorphisms (SNPs) rs7412 and rs429358 downloaded from the genotyped data were used to determine the APOE genotype ^27^. APOE was considered as the major genetic AD risk factor, however, it was also reported to be associated with cerebrovascular lesions. As a result, APOE ε4 carrier status was also included in this study as a covariate and was classified into three categories based on the number of ε4 alleles, i.e., non-carriers; carriers with one ε4 allele; carriers with two ε4 alleles.

### 2.5 Cognitive tests

Seven neuropsychological tests were included in this study: Reaction Time, Trail Making Test A and Symbol Digit Substitution for assessing *processing speed*; Numeric Memory and Pairs Matching for assessing *memory*; Trail Making Test B and Fluid Intelligence for assessing *executive function*. Raw scores for these tests were standardised by transforming the raw cognitive scores into z-scores using the healthy reference subsample of UK Biobank at baseline. Specific cognitive domain scores were computed by averaging the corresponding cognitive test scores within a domain and then standardising against the healthy subsample. Global cognition was computed by averaging the scores across all three cognitive domains and again standardising against the healthy subsample. Further details of the standardisation procedure can be found in our previous work using the UK Biobank data ^28^.

### 2.6 Statistical analysis

Statistical analyses were conducted using SPSS (IBM corporation, USA) version 26.0 and R version 3.6.1. Two-tailed p < 0.05 was considered statistically significant. The difference of WMBAG between healthy and unhealthy participants in the baseline test set was compared using Analysis of Covariance (ANCOVA) adjusting for chronological age, sex, scanner and APOE status. Comparison of baseline age, sex, education and risk factors between test participants who had only one time-point brain scan and 1409 participants who had repeat (follow-up) scans was conducted using independent-t test and χ^2^ analysis, while comparison of white matter brain age measures and cognition was performed by ANCOVA controlling for baseline chronological age, sex, and scanner and APOE.

Multiple linear regression models were conducted to investigate the associations between vascular risk factors and WMBAG at baseline. VRS was dummy coded with the 0 category as reference in all analyses. Eight regression models were used in this analysis for analysing the vascular risk factors -see their mathematical expressions in Supplementary methods. For model 1a, we investigated the main effect of VRS on the white matter brain age gap. For model 1b, interaction terms (sex times different VRS levels) were added to the model to examine if the effects of VRS differ by sex. In model 2a, to determine the specific contribution of each risk factor, the VRS was replaced with all five vascular risk factors. In model 2b-f, we investigated the moderation effect of sex on the relationship between each vascular risk factor and WMBAG. Chronological age, sex, scanner and APOE status were controlled for all models.

The association between WMBAG and cognition at baseline was first examined. Then mediation analysis with WMBAG as a mediator and cognition as outcome, was carried out among baseline participants with VRS and individual risk factors as predictors. Baseline chronological age, sex, scanner, APOE and education were controlled. Bonferroni correction was applied for these analyses with four cognitive outcomes (corrected alpha level = 0.05/4 = 0.0125). Mediation analysis was performed using the ‘mediation’ package^29^ in R. Direct and indirect effects were estimated via bootstrapping with 5,000 samples.

For longitudinal analysis, a dependent t-test was conducted to examine change in WMBAG between baseline and follow-up. To explore the prospective effects in the longitudinal subset, we first conducted multiple linear regression to examine the relationships between baseline vascular risk factors and change in WMBAG (calculated as the difference between follow-up and baseline scores), and that between WMBAG change and cognition change. Mediation analysis was then conducted to examine the direct and indirect effects of vascular risk factors on change in cognition, through change in WMBAG.

### 2.7 Data and code availability statement

The UK Biobank data can be accessed by online application (https://www.ukbiobank.ac.uk/). Codes for the 3D-CNN deep learning model in this study can be shared from the authors upon request.

## 3. RESULTS

### 3.1 Sample characteristics

Sample characteristics including demographics and vascular risk factors of test data are shown in Table 1. Cross-sectional test data included 7769 healthy participants and 3399 unhealthy participants. Among them, 1409 participants with both the baseline and follow-up scans were used for longitudinal analysis.

**Table 1.**
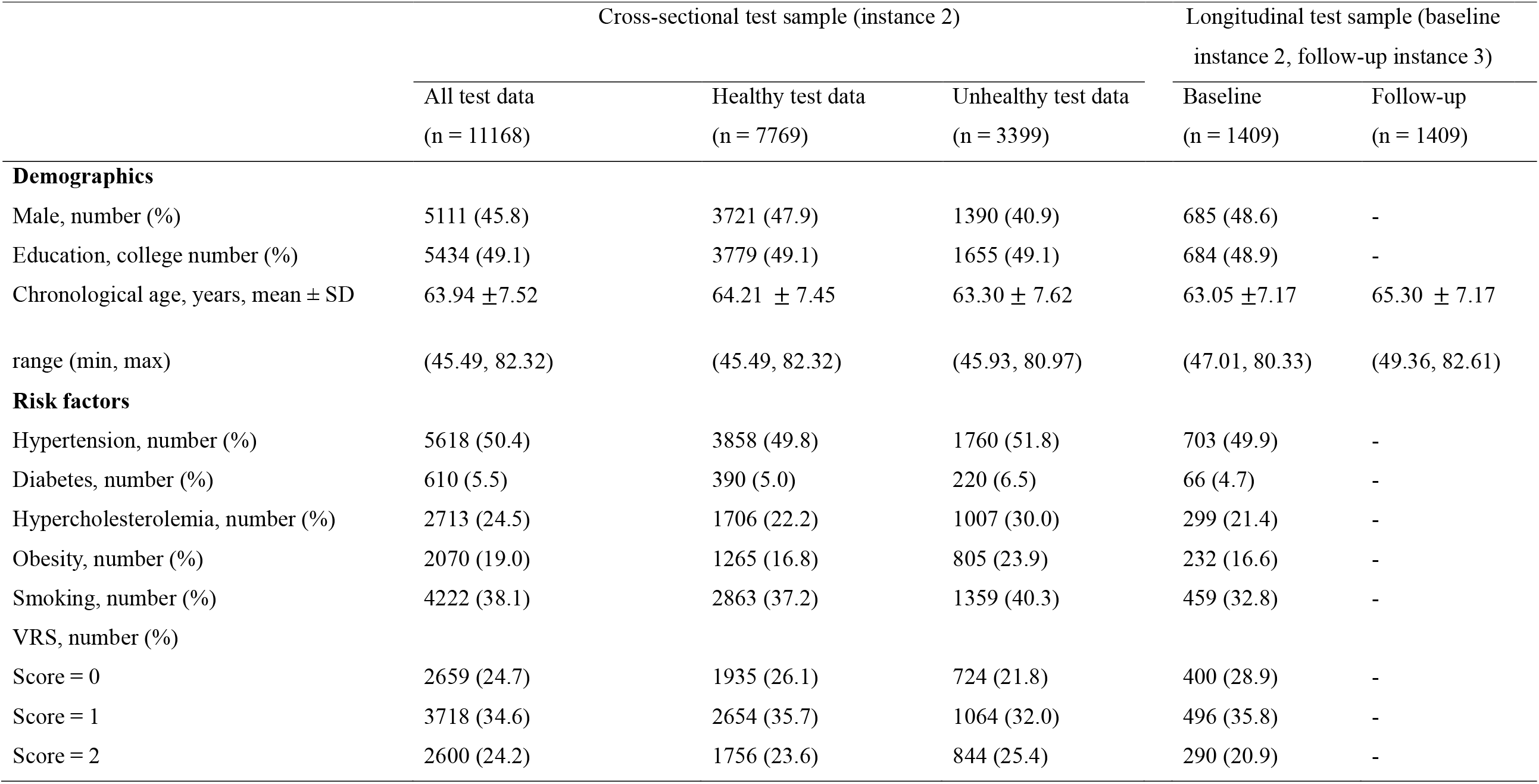
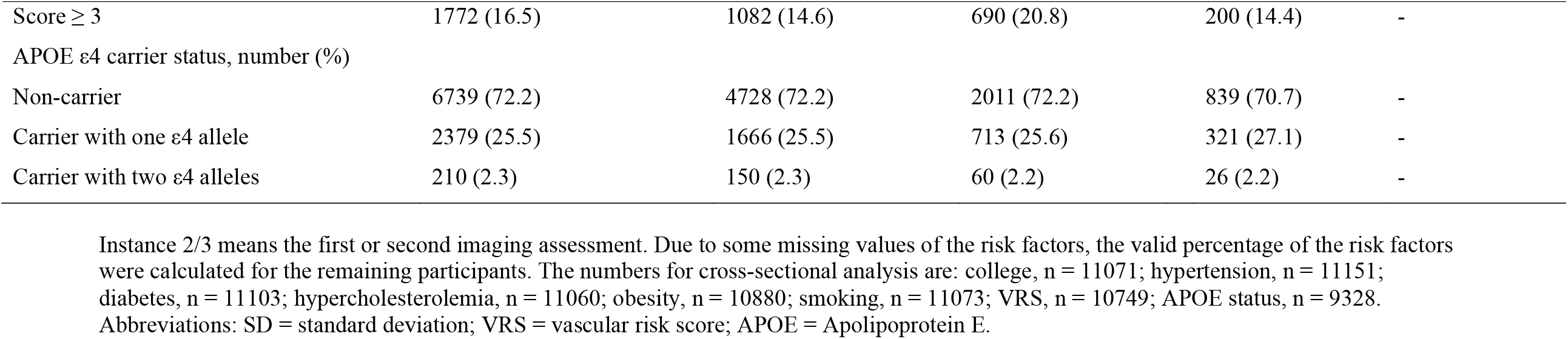
Characteristics of test samples.

### 3.2 White matter brain age prediction

The white matter brain age predictions before and after bias correction for the whole test set are shown in Figure 3 (A and B). Spearman correlation coefficient between WMBAG and chronological age for the whole cross-sectional test participants was reduced from -0.54 before bias correction to 0.04 after bias correction, with a slight increase of MAE from 2.57 to 2.84. Pearson’s r between white matter brain age and chronological age is 0.902.

**Figure 3.**
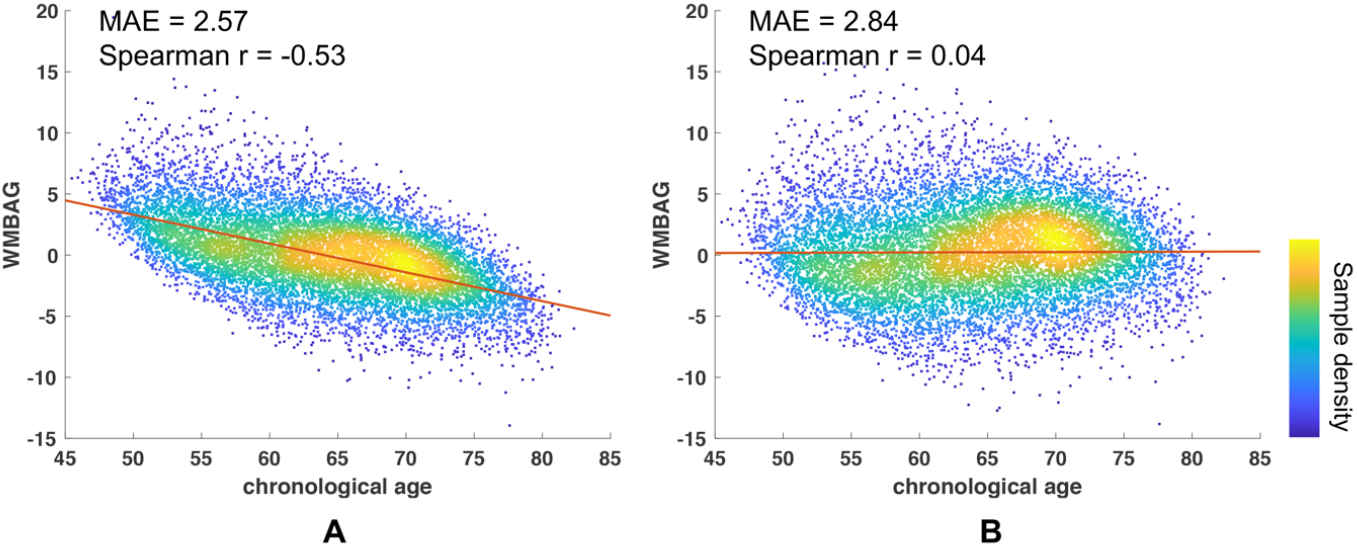
Bias correction. Association between chronological age and uncorrected WMBAG (A); Association between chronological age and bias-corrected WMBAG (B). All cross-sectional test participants were included in this bias correction analysis (n= 11168). This bias correction was conducted using the predicted age after fusion of five DWI-derived maps. MAE and the correlation coefficient (r) were listed in the upper left corner of each sub-plot. Colour bar indicates the sample density. WMBAG = White matter brain age – Chronological age. Abbreviations: WMBAG = white matter brain age gap; Spearman r = coefficient for Spearman correlation; MAE = mean absolute error; WM = white matter; DWI = diffusion weighted maps.

Cross-sectional white matter brain age which was computed using our 3D-CNN model and WMBAG are summarised in Table 2. Interestingly, participants with none of the vascular risk factors had a negative mean WMBAG of 0.56, which suggested that they had a brain 0.56 years younger on average than their chronological age. Performances for all DWI maps and fusion results are listed in Table 3. The MAE for the fused white matter brain age was smaller than that of any other single DWI derived map. MAE measured on the healthy test data was 2.75 years (Table 3 and Supplementary Figure e-1A) with Pearson’s r between chronological and predicted brain age of 0.908 (p < 0.001). For unhealthy test data (Table 3 and Supplementary Figure e-1B), the MAE was 3.03 with Pearson’s r = 0.892 (p < 0.001). Due to the best performance of the fusion of all five DWI maps, we conducted the subsequent clinical analysis using the fused predicted age. The mean WMBAG for unhealthy test participants was 0.51 ± 0.08 years older than healthy test participants (p < 0.001, 95% CI =0.348 – 0.668). Comparison of baseline characteristics between 9759 baseline participants with single time-point scans and 1409 participants who had repeat scans were listed in Supplementary Table e2; at baseline, the WMBAG of the participants who had follow-up brain scans were 0.36 ± 0.11 years lower than that of the rest baseline participants. The cognitive scores such as processing speed, executive function, memory, and global cognition of the participants with follow-up scans were significantly higher than those of the rest of the test sample participants at baseline (see supplementary Table e2).

**Table 2.** White matter brain age computed using 3D-CNN and WMBAG.

| | White matter brain age, years, mean ± SD<br>range (min, max) | WMBAG, years, mean ± SD<br>range (min, max) |
| --- | --- | --- |
| **Cross-sectional test sample (instance 2)** |  |  |
| All (n = 11168) | 64.17 ± 8.35 (45.13, 84.06) | 0.23 ± 3.60 (-13.82, 20.91) |
| Healthy test data (n = 7769) | 64.30 ± 8.31 (45.13, 83.60) | 0.09 ± 3.49 (-12.50, 13.94) |
| Unhealthy test data (n = 3399) | 63.86 ± 8.45 (45.73, 84.06) | 0.56 ± 3.82 (-13.82, 20.91) |
| **Longitudinal test sample (baseline instance 2, follow-up instance 3)** |  |  |
| Baseline (n = 1409) | 62.94 ± 8.02 (45.71, 81.77) | -0.11 ± 3.45 (-12.25, 13.59) |
| Follow-up (n = 1409) | 65.28 ± 8.05 (46.02, 82.98) | -0.02 ± 3.45 (-12.46, 12.90) |
| **VRS levels for all test data** |  |  |
| Score = 0 (n = 2659) | 60.57 ± 7.91 (45.13, 82.23) | -0.56 ± 3.53 (-12.06, 20.91) |
| Score = 1 (n = 3718) | 63.30 ± 8.15 (45.73, 83.22) | -0.02 ± 3.55 (-12.72, 15.70) |
| Score = 2 (n = 2600) | 66.16 ± 7.88 (46.26, 84.06) | 0.64 ± 3.54 (-13.82, 14.68) |
| Score ≥ 3 (n = 1772) | 68.19 ± 7.41 (47.01, 83.60) | 1.29 ± 3.58 (-10.94, 14.79) |

Instance 2/3 means the first or second imaging assessment. Abbreviations: 3D-CNN = three-dimensional convolutional neural network; WMBAG = white matter brain age gap; SD = standard deviation; VRS = vascular risk score.

**Table 3.** Model performance for different DWI maps.

|  | Healthy (n = 7769) |  |  |  | Unhealthy (n = 3399) |  |  |  |
| --- | --- | --- | --- | --- | --- | --- | --- | --- |
|  | Before bias correction |  | After bias correction |  | Before bias correction |  | After bias correction |  |
|  | MAE (years) | Pearson's r | MAE (years) | Pearson's r | MAE (years) | Pearson's r | MAE (years) | Pearson's r |
| FA | 2.76 | 0.890 | 3.05 | 0.890 | 2.95 | 0.882 | 3.24 | 0.882 |
| MD | 2.77 | 0.884 | 3.16 | 0.884 | 3.05 | 0.865 | 3.45 | 0.865 |
| AxD | 2.78 | 0.886 | 3.16 | 0.888 | 3.07 | 0.868 | 3.43 | 0.868 |
| RD | 2.73 | 0.886 | 3.13 | 0.886 | 2.99 | 0.867 | 3.44 | 0.867 |
| MO | 3.01 | 0.875 | 3.53 | 0.875 | 3.21 | 0.848 | 3.74 | 0.848 |
| fusion | 2.51 | 0.908 | 2.75 | 0.908 | 2.71 | 0.892 | 3.03 | 0.892 |

This table shows the model performance in healthy and unhealthy test data before and after bias correction. Abbreviations: DWI = diffusion weighted imaging; FA = fractional anisotropy; MD = mean diffusivity; AxD = axial diffusivity; RD = radial diffusivity; MO = anisotropy mode; MAE = mean absolute error; fusion = fusion of all five diffusion weighted maps; Pearson's r = Pearson's correlation coefficient.

### 3.3 Cross-sectional analysis at baseline

#### 3.3.1 Associations between risk factors and WMBAG

In model 1a, after controlling for chronological age, sex, scanner and APOE status, participants with one, two, and three or more vascular risk factors had an increased WMBAG of 0.54, 1.23, and 1.94 years older, respectively, than those without vascular risk factors (Table 4; also see Figure 4A). In model 1b, significant interaction between sex and VRS on its association with WMBAG was found (p = 0.015; Table 4). Among participants with three or more vascular risk factors, the WMBAG of males was significantly larger than that of females (mean difference = 0.617 years, p = 0.001). No significant difference of WMBAG between males and females was found for those with 0, 1 or 2 risk factors (Figure 4B).

**Table 4.** Association between VRS and WMBAG.

|  |  | Unstandardised beta | 95% CI |  | p-value |
| --- | --- | --- | --- | --- | --- |
|  |  |  | Lower bound | Upper bound |  |
| Main effect<br>(model 1a) | Chronological age | -0.026 | -0.036 | -0.016 | < 0.001 |
|  | Sex | 0.087 | -0.063 | 0.237 | 0.258 |
|  | Scanner | 0.231 | 0.145 | 0.317 | < 0.001 |
|  | APOE status | 0.047 | -0.098 | 0.192 | 0.528 |
|  | VRS_1 | 0.538 | 0.345 | 0.730 | < 0.001 |
|  | VRS_2 | 1.229 | 1.014 | 1.444 | < 0.001 |
|  | VRS_3 | 1.936 | 1.692 | 2.181 | < 0.001 |
| Interactions<br>(model 1b) | Chronological age | -0.026 | -0.036 | -0.016 | < 0.001 |
|  | Sex | 0.01 | -0.299 | 0.318 | 0.951 |
|  | Scanner | 0.232 | 0.146 | 0.318 | < 0.001 |
|  | APOE status | 0.049 | -0.096 | 0.194 | 0.507 |
|  | VRS_1 | 0.543 | 0.301 | 0.785 | < 0.001 |
|  | VRS_2 | 1.282 | 1.000 | 1.564 | < 0.001 |
|  | VRS_3 | 1.572 | 1.217 | 1.928 | < 0.001 |
|  | VRS_1*Sex | 0.002 | -0.395 | 0.400 | 0.991 |
|  | VRS_2*Sex | -0.076 | -0.505 | 0.354 | 0.730 |
|  | VRS_3*Sex | 0.607 | 0.119 | 1.095 | 0.015 |

Main effect of VRS on WMBAG was analysed by recoding VRS into dummy variables as independent variables. Interaction effects were analysed by adding their corresponding interaction terms to the model. Abbreviations: WMBAG = white matter brain age gap; VRS = vascular risk score; APOE = Apolipoprotein E; CI = confidence interval, VRS\_1 is the dummy variable indicating participants with only 1 vascular risk factor; VRS\_2 indicates participants with 2 vascular risk factors; VRS\_3 indicates participants with 3 or more vascular risk factors.

**Figure 4.**
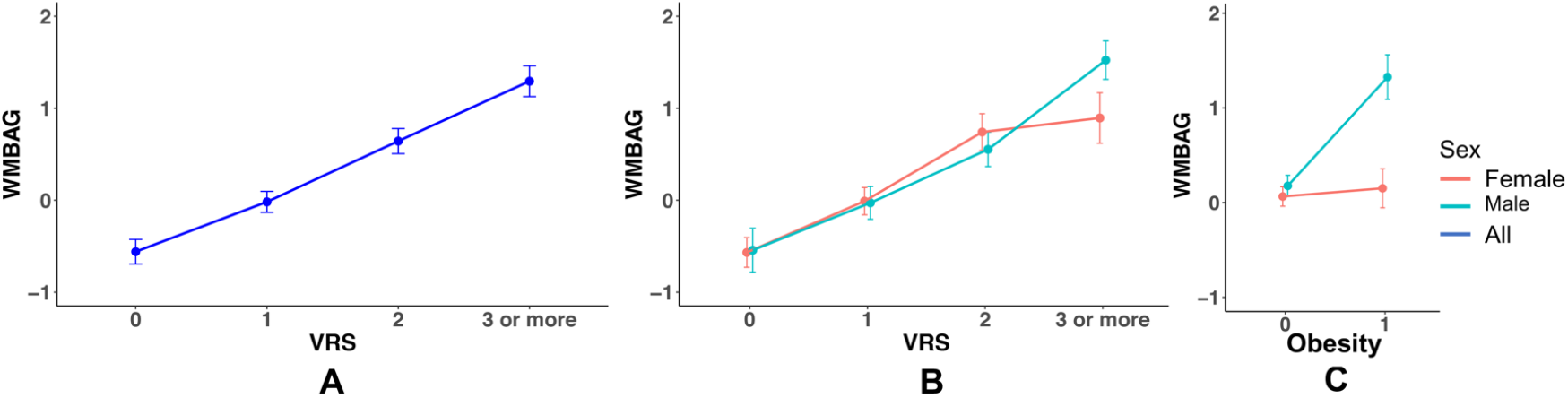
Difference of WMBAG across different VRS groups (A for all participants; B for males and females separately) and WMBAG difference for different obesity status by sex (C). Each dot indicated the mean value for the WMBAG, error bar indicated the 95% CI. Abbreviations: WMBAG = white matter brain age gap; VRS = vascular risk score; WM = white matter. CI = confidence interval.

Apart from the composite VRS, we also observed significant unique contributions of each individual risk factor (except obesity) to the WMBAG when controlling for other risk factors and covariates, in model 2a (Table 5). Having diabetes was more strongly associated with WMBAG (1.39 years, p = 0.002), relative to other risk factors. Interestingly, in models 2b-f, we found that all interaction terms were not significant, except for the interaction between obesity and sex (Table 5). We found that female participants with obesity did not have larger WMBAG, while males with obesity had significant larger WMBAG than those without obesity (Figure 4C). Different APOE ε4 carrier status was not associated with the WMBAG in any of the models (all p values > 0.05).

**Table 5.** Associations between different vascular risk factors and WMBAG.

|  |  | Unstandardised beta | 95%CI |  | p-value |
| --- | --- | --- | --- | --- | --- |
|  |  |  | Lower bound | Upper bound |  |
| Main effects<br>(model 2a) | Chronological age | -0.028 | -0.038 | -0.017 | < 0.001 |
|  | Sex | 0.052 | -0.099 | 0.202 | 0.503 |
|  | Scanner | 0.235 | 0.149 | 0.321 | < 0.001 |
|  | APOE status | 0.057 | -0.088 | 0.202 | 0.440 |
|  | Hypertension | 0.871 | 0.713 | 1.028 | < 0.001 |
|  | Diabetes | 1.390 | 0.119 | 0.503 | 0.002 |
|  | Hypercholesterolemia | 0.311 | 1.051 | 1.729 | < 0.001 |
|  | Obesity | 0.161 | -0.033 | 0.355 | 0.103 |
|  | Smoking | 0.689 | 0.537 | 0.841 | < 0.001 |
| Interactions<br>(models 2b-f) | Hypertension *Sex | 0.112 | -0.183 | 0.406 | 0.458 |
|  | Diabetes*Sex | 0.318 | -0.278 | 1.040 | 0.257 |
|  | Hypercholesterolemia*Sex | -0.221 | -0.574 | 0.131 | 0.219 |
|  | Obesity*Sex | 1.023 | 0.643 | 1.403 | < 0.001 |
|  | Smoking*Sex | 0.275 | -0.026 | 0.576 | 0.073 |

Independent main effects of vascular risk factors on WMBAG were analysed by adding all vascular risk factors into the regression model. Interaction effects were analysed by adding each vascular risk factor and its corresponding interaction term to the model. Abbreviations: WMBAG = white matter brain age gap; APOE = Apolipoprotein E; CI = confidence interval.

#### 3.3.2 Association between cognition and WMBAG

After controlling for age, sex, scanner, APOE and education, WMBAG was found to be significantly and negatively associated with baseline processing speed (unstandardised b = - 0.025, p < 0.001), executive function (unstandardised b = -0.018, p < 0.001), memory (unstandardised b = -0.008, p = 0.027) and global cognition (unstandardised b = -0.022, p < 0.001). However, only speed, executive function and global cognition survived after the Bonferroni correction.

Mediation analysis on baseline data is summarized in Table 6. Significant mediation effects of WMBAG were observed for the associations between hypertension and processing speed, executive function and global cognition (bs = -0.019 ∼ -0.014, all p values < 0.001), and the associations between diabetes and these cognitive outcomes (bs = -0.033 ∼ -0.024, all p values < 0.001). Smoking was also found to be associated with processing speed and executive function decline via WMBAG (b = -0.014, p < 0.001 and b = -0.010, p < 0.001, respectively), and was also associated with executive function, memory, and global cognition directly (bs = -0.071 ∼ -0.007, all p values < 0.05. Obesity was associated with processing speed decline directly (b = -0.103, p = 0.001), but not mediated by WMBAG (b = 0.003, p = 0.200).

**Table 6.** Associations between baseline vascular risk factors and cognition mediated by WMBAG.

|  | Processing speed |  |  |  | Executive |  |  |  | Memory |  |  |  | Global cognition |  |  |  |
| --- | --- | --- | --- | --- | --- | --- | --- | --- | --- | --- | --- | --- | --- | --- | --- | --- |
|  | Unstandardised beta | 95%CI |  | p-value | Unstandardised beta | 95%CI |  | p-value | Unstandardised beta | 95%CI |  | p-value | Unstandardised beta | 95%CI |  | p-value |
|  |  | Lower bound | Upper bound |  |  | Lower bound | Upper bound |  |  | Lower bound | Upper bound |  |  | Lower bound | Upper bound |  |
| **Direct effects** |  |  |  |  |  |  |  |  |  |  |  |  |  |  |  |  |
| Hypertension | -0.020 | -0.078 | 0.003 | 0.400 | 0.011 | -0.041 | 0.060 | 0.690 | -0.026 | -0.081 | 0.030 | 0.330 | -0.015 | -0.063 | 0.030 | 0.550 |
| Diabetes | -0.018 | -0.132 | 0.100 | 0.750 | -0.035 | -0.160 | 0.090 | 0.590 | -0.070 | -0.197 | 0.060 | 0.290 | -0.051 | -0.175 | 0.070 | 0.400 |
| Hypercholesterolemia | -0.043 | -0.099 | 0.010 | 0.148 | -0.051 | -0.114 | 0.010 | 0.094 | -0.028 | -0.094 | 0.040 | 0.420 | -0.051 | -0.108 | 0.010 | 0.078 |
| Obesity | **-0.103** | -0.162 | -0.050 | 0.001 | -0.059 | -0.123 | 0.010 | 0.082 | -0.027 | -0.040 | 0.090 | 0.460 | -0.056 | -0.116 | 0.000 | 0.066 |
| Smoking | -0.028 | -0.073 | 0.020 | 0.218 | **-0.071** | -0.121 | -0.020 | 0.004 | **-0.071** | -0.122 | -0.020 | 0.005 | **-0.007** | -0.012 | -0.020 | 0.004 |
| **Indirect effects mediated through baseline WMBAG** |  |  |  |  |  |  |  |  |  |  |  |  |  |  |  |  |
| Hypertension | **-0.019** | -0.026 | -0.001 | < 0.001 | **-0.014** | -0.021 | -0.010 | < 0.001 | -0.005 | -0.012 | 0.000 | 0.140 | **-0.016** | -0.023 | -0.010 | <0.001 |
| Diabetes | **-0.033** | -0.048 | -0.002 | < 0.001 | **-0.024** | -0.038 | -0.010 | < 0.001 | -0.009 | -0.022 | 0.000 | 0.130 | **-0.027** | -0.042 | -0.020 | <0.001 |
| Hypercholesterolemia | -0.004 | -0.009 | 0.000 | 0.094 | -0.003 | -0.007 | 0.000 | 0.096 | -0.001 | -0.004 | 0.000 | 0.200 | -0.003 | -0.008 | 0.000 | 0.108 |
| Obesity | 0.003 | -0.090 | 0.000 | 0.200 | -0.002 | -0.007 | 0.000 | 0.210 | -0.0009 | -0.003 | 0.000 | 0.310 | -0.003 | -0.008 | 0.000 | 0.194 |
| Smoking | **-0.014** | -0.021 | -0.010 | < 0.001 | **-0.010** | -0.017 | -0.010 | < 0.001 | -0.0001 | -0.001 | 0.000 | 0.758 | 0.0001 | 0.000 | 0.000 | 0.036 |

This table shows the mediation effect of baseline WMBAG on the associations between baseline vascular risk factors and baseline cognition. Chronological age, sex, scanner, APOE and education were controlled for all models. Raw p values are reported in this table with bold unstandardised beta indicating statistical significance after Bonferroni correction (corrected $\alpha$ level = 0.0125). Abbreviations: WMBAG = white matter brain age gap; CI = confidence interval.

### 3.4 Longitudinal analysis

The demographics of 1409 participants with baseline and follow-up scans are shown in Table 1. Estimated white matter brain age and WMBAG for both timepoints are presented in Table 2. Generally, participants underwent an average 2.25 ± 0.12 years of follow up (ranging from 2.01 to 2.67 years). One thousand three hundred and fourteen (93.26%) participants had increased white matter brain age with an average of 2.57 ± 1.48 years (dependent t-test, p < 0.001) between baseline and follow-up scans (Figure 5). VRS was not associated with the WMBAG change, no individual vascular risk factors contributed significantly to the WMBAG change expect for obesity (Supplementary Table e-3). While the presence of obesity contributed to decrease in WMBAG (unstandardised b = -0.371, p = 0.008), the direction of which was not within our expectation. No significant associations between WMBAG change and cognition change were observed (Supplementary Table e-4). Moreover, we did not find any significant mediation effect of WMBAG change on the relationships between vascular risk factors and cognition change (all p values > 0.05, see Supplementary Table e-5).

**Figure 5.**
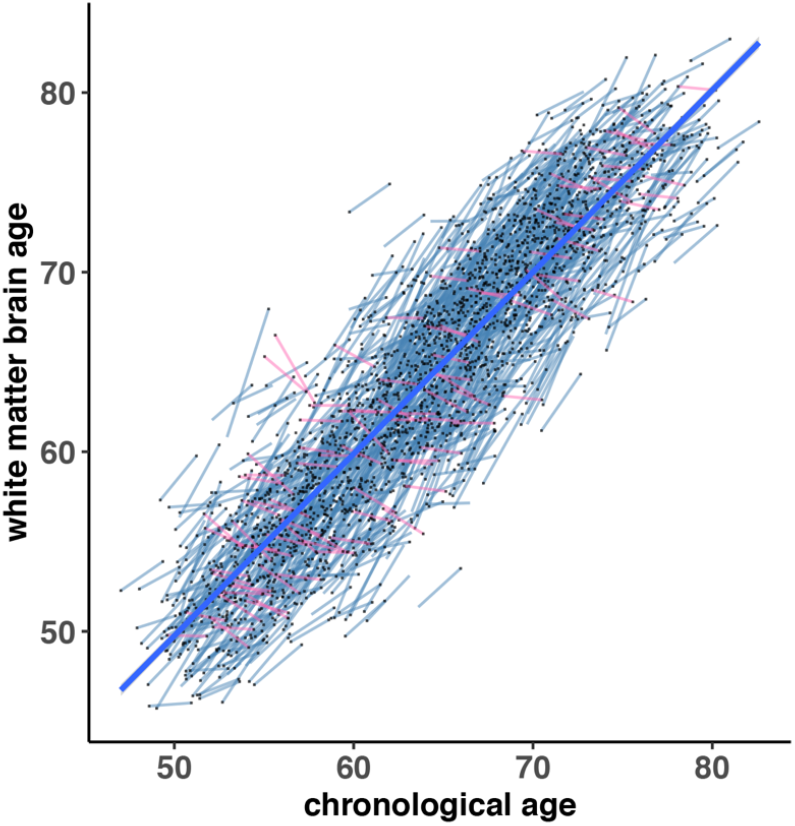
Longitudinal change of white matter brain age for each participant. Each participant has two time-point white matter brain ages (shown as two dots), connected with either a short blue line indicating white matter brain age increase (93.26%), or a red line indicating white matter brain age decrease. The bold blue line was a mean line fitted using white matter brain ages from all participants at both two timepoints.

## 4. DISCUSSION

This study had three main findings. First, we successfully developed a deep learning model using convolutional neural network (CNN) for estimating brain age based on cerebral white matter only. Our model produced robust and accurate white matter brain age prediction in healthy test subjects with MAE of 2.75 and Pearson’s r of 0.908 with chronological age. Second, we found that, cross-sectionally, cerebrovascular risk factors, both individually and collectively, were significantly associated with WMBAG, such that participants free of vascular risk factor had a younger brain (WMBAG = -0.56); higher WMBAG was associated with poorer cognitive performance, especially processing speed and executive function. Third, we demonstrated that WMBAG played a mediation role between vascular risk factors, namely, hypertension and diabetes, and declined cognition, especially with a slower processing speed and worse executive function.

Our trained 3D-CNN model showed a better MAE in age prediction compared with many previous brain age studies^30^. Technically, this model was trained in a large population sample of healthy community-dwelling participants drawn from the UK Biobank, which enabled strong power for model estimation. The combined information extracted from all five DWI maps also improved the model accuracy; as shown in Table 3, the information from the fusion of five DWI maps resulted in a lower MAE and higher Pearson’s coefficient. FA, MD, AxD, RD and MO maps are widely recognized DWI feature maps and have been shown to be significantly correlated with vascular risk factors^31, 32^. Each of them taps into distinct physiological properties of the white matter microstructure, from which the deep learning model extracted essential information for white matter brain age prediction. We conducted the bias correction for the original white matter brain age to remove the negative bias towards the chronological age. Taking all these technical steps into consideration, our deep learning model for white matter age predication was well established. The mean WMBAG for unhealthy subjects was shown to be 0.51 ± 0.08 years greater than that of those healthy subjects. The 1409 participants who had repeat brain MRI scans did not only have better cognition, but also had a 0.36 ± 0.11 years younger brain age than the participants who had only a single time-point scan (p < 0.001). This might suggest the potential recruiting bias, as it was reported that the response rate to the repeat MRI scan was 65%^33^. Our method’s ability to detect the subtle brain age difference between the two groups of participants from the same cohort with only sample recruitment difference shows that our brain age estimation was accurate and reliable. Moreover, we also validated the model’s performance in the subsample with two time-point scans. After approximately an average of 2.25 years of follow up, 93.26% of the 1409 participants had an increased white matter brain age compared with baseline, which further demonstrated that our deep learning model was robust and able to extract useful information from the DWI maps for white matter brain age prediction.

Interestingly, the WMBAG computed in this study correlated with the cerebrovascular burden but not with the neurodegenerative risk factor, APOE genotype. Neurodegenerative and vascular risk factors are both associated with brain ageing^34, 35^. In our study, except for obesity, all vascular risk factors were significantly correlated with WMBAG with diabetes and hypertension having the highest correlations. Our results suggested that the diabetic participants on average had a WMBAG of 1.39-years older than that of the non-diabetic participants; similarly, the brain of a hypertensive participant would have a WMBAG of 0.871 years older than those without hypertension. Collectively, the accumulation of vascular risk factors led to a larger WMBAG, suggesting that WMBAG is a sensitive biomarker for monitoring the vascular burden. This finding was consistent with previous observations that hypertension and diabetes are the most widely recognized vascular risk factors that are highly associated with morbidity and mortality of cerebrovascular diseases (CVD)^16, 36-38^. One previous study also reported that DWI based brain age gap was correlated with blood pressure and smoking^39^. However, no significant association was found between the APOE ε4 allele(s) status and WMBAG. APOE ε4 allele(s) has been recognized as the strongest genetic risk factor for sporadic Alzheimer’s Disease^40^. APOE genotypes with one or two ε4 allele(s) lead to a three to 10 fold risk for AD, respectively^41^. While some studies also reported that AOPE was correlated with subcortical lesions such as WMH^42^ and microbleeds^43^, the findings were not always consistent. Most previous brain age studies aimed at capturing the overall changes for the whole brain, therefore unable to differentiate cerebrovascular burden from neurodegenerative burden. Although increasing evidence has suggested cerebrovascular disease and neurodegenerative disease share multiple risk factors and have overlapping neuropathologies^44, 45^, there is a general difference in their MRI manifestations. AD patients usually start grey matter atrophy at premorbid stage and the atrophy progresses with the advance of AD, while those with cerebrovascular disease usually suffer more from the subcortical lesions such as WMH, lacunes, microbleeds and enlarged perivascular spaces^13^. Our study was based on this hypothesis, and we used DWI for white matter brain age computation, given its sensitivity to the microstructural integrity and pathology of subcortical white matter.

Sex dimorphism was observed when we considered the interactive effect between sex and vascular risk factors on the prediction of the WMBAG. Males showed higher WMBAG than females when they had three or more vascular risk factors. Obesity was the only vascular risk factor that showed interactive effect with sex on the WMBAG. Only males with obesity had a significantly greater WMBAG, suggesting that obesity was detrimental to brain ageing in men but not in women. This finding was in line with the finding of our previous study which investigated sex difference in WMH^46^; males with higher BMI showed significantly greater deep WMH volume compared to women. Although the potential mechanisms underlying this sex difference have not been fully understood, other studies have also reported this interesting dimorphism, suggesting that females might be more resilient to the detrimental effect of obesity on the brain than males^47^. A cross-sectional study conducted in the UK Biobank cohort also found that obese men had a steeper grey matter volume decline than women^48^. One hypothesis posits that the distribution of adipose tissues in males and females is different - males tend to accrue more visceral fat, which heightens the vascular burden; conversely women usually accrue more fat in the subcutaneous depot, which is an independent predictor of lower cardiovascular and diabetes-related mortality^49^.

Significant associations between WMBAG processing speed, executive function and global cognition after Bonferroni correction were observed cross-sectionally. Processing speed and executive function were considered to be the most vulnerable cognitive domains in CVD^50^. In comparison with AD patients, patients with CVD usually show less pronounced memory deficits^51^, although the memory dysfunction may also appear progressively during the later course of the disease. Consistent with the clinical differentiations between AD and CVD, we did not find a significant association between the WMBAG and memory loss after Bonferroni correction, which further demonstrated both specificity and reliability of our white matter brain age model in relation to the cerebrovascular disease burden, and that our model may have clinical utility.

Using mediation analyses in baseline participants, we found that among the five cerebrovascular risk factors, only hypertension and diabetes were associated with processing speed, executive function, and global cognition through the mediation of WMBAG. These findings validated the underlying pathway that the vascular risk factors would contribute to the pathological changes in the white matter and then lead to the cognitive dysfunction. However, obesity was the only vascular risk factor in our study that contributed directly to the cognitive decline but not through the WMBAG. Although some studies have observed a mediation effect of white matter changes on the relationship between obesity and cognition in healthy adults^52^, our result suggested otherwise. These modifiable vascular risk factors were also reported to be linked to late-life progression of brain disorders in longitudinal studies^53, 54^, but few studies have conducted the mediation analysis in a longitudinal cohort with a relatively large sample size. However, our longitudinal analysis did not yield a significant mediation effect of WMBAG change between any vascular risk factor and cognitive decline. This may be partly due to the short period of time between baseline and follow-up (i.e., about 2 years), where significant changes in WMBAG might be too subtle to be detected.

Moreover, many participants had better cognition at follow-up than baseline due perhaps to practice effects (data not shown).

We believe that future work should be carried out to further investigate the relationship between vascular risk factors and white matter brain age. Our stratification for the level of risk factors was based on the number of vascular risk factors, regardless of the type of vascular risk factors a participant had or the specific contribution of each risk factor. For example, a participant with diabetes only would be grouped with anyone with just one of the vascular risk factors we investigated regardless of the type, i.e., any of one of hypertension, diabetes, hypercholesterolemia, obesity or smoking. In this study, we ‘binarized’ our participants into ‘presence’ or ‘absence’ of a vascular risk factor. Comparisons were therefore limited to ‘yes’ or ‘no’ as to whether the participant had that particular vascular risk factor or not, with a lack of more nuanced investigations of the disease stage or disease severity dependent effects of clinical measurements on these risk factors. Additionally, although we have a longitudinal subset with a large sample size from UK Biobank, the follow-up time might be too short to uncover significant brain structural and cognitive changes. Some cerebrovascular and neurodegenerative pathologies may coexist in the brain ageing process, and it is difficult to differentiate the effect of these pathologies on white matter and grey matter distinctively.

## Data Availability

The UK Biobank data used in this study can be accessed by online application (https://www.ukbiobank.ac.uk/). Codes for the 3D-CNN deep learning model in this study can be shared from the authors upon request.

## 5. ACKNOWLEDGEMENTS

This research was conducted under the Application ID 45262 and 37103; This research was undertaken with the assistance of resources and services from the National Computational Infrastructure (NCI), which is supported by the Australian Government. Ivor W Tsang and Yuangang Pan were supported by the A*STAR Centre for Frontier AI Research. We also thank Angie Russell for her assistance in the preparation of the manuscript.

## 6. AUTHOR CONTRIBUTION

Writing – JD, YP, PSS, JJ, AT, WW, Conception of idea – JD, WW, JJ. Computation and coding– YP, IWT, JJ. Statistics/Analysis – JD, BL, RC, AT. Cognition – JD, BL. Comments/edits – all.

## 7. DECLARATION OF COMPETING INTEREST

The authors declare no conflict of interest.

## 8. STUDY FUNDINGS

This study was supported by the National Health and Medical Research Council (NHMRC) of Australia Program Grants ID350833, ID568969, ID1093083 and ID630593; Jiyang Jiang was supported by John Holden Family Foundation.

## Notes

### Competing Interest Statement

The authors have declared no competing interest.

### Author Declarations

This study involves only openly available human data, which can be obtained from:https://www.ukbiobank.ac.uk/

## REFERENCE

1. Peters R. Ageing and the brain. Postgrad Med J. 2006;82(964):84–8.

2. Cole JH, Marioni RE, Harris SE, Deary IJ. Brain age and other bodily ‘ages’: implications for neuropsychiatry. Mol Psychiatry. 2019 Feb;24(2):266–81.

3. Cole JH, Franke K. Predicting Age Using Neuroimaging: Innovative Brain Ageing Biomarkers. Trends Neurosci. 2017 Dec;40(12):681–90.

4. Jonsson BA, Bjornsdottir G, Thorgeirsson TE, et al. Brain age prediction using deep learning uncovers associated sequence variants. Nat Commun. 2019 Nov 27;10(1):5409.

5. Ly M, Yu GZ, Karim HT, et al. Improving brain age prediction models: incorporation of amyloid status in Alzheimer’s disease. Neurobiol Aging. 2020 Mar;87:44–8.

6. Hajek T, Franke K, Kolenic M, et al. Brain Age in Early Stages of Bipolar Disorders or Schizophrenia. Schizophr Bull. 2019 Jan 1;45(1):190–8.

7. Beheshti I, Mishra S, Sone D, Khanna P, Matsuda H. T1-weighted MRI-driven Brain Age Estimation in Alzheimer’s Disease and Parkinson’s Disease. Aging Dis. 2020 May;11(3):618–28.

8. Cole JH, Raffel J, Friede T, et al. Longitudinal Assessment of Multiple Sclerosis with the Brain-Age Paradigm. Ann Neurol. 2020 Jul;88(1):93–105.

9. Ge Y, Grossman RI, Babb JS, Rabin ML, Mannon LJ, Kolson DL. Age-related total gray matter and white matter changes in normal adult brain. Part I: volumetric MR imaging analysis. AJNR Am J Neuroradiol. 2002 Sep;23(8):1327–33.

10. Bose SK, Mackinnon T, Mehta MA, et al. The effect of ageing on grey and white matter reductions in schizophrenia. Schizophr Res. 2009 Jul;112(1-3):7–13.

11. Iadecola C. The pathobiology of vascular dementia. Neuron. 2013 Nov 20;80(4):844–66.

12. Agarwal N, Carare RO. Cerebral Vessels: An Overview of Anatomy, Physiology, and Role in the Drainage of Fluids and Solutes. Frontiers in Neurology. 2021 2021-January-13;11(1748).

13. Wardlaw JM, Smith EE, Biessels GJ, et al. Neuroimaging standards for research into small vessel disease and its contribution to ageing and neurodegeneration. Lancet Neurol. 2013 Aug;12(8):822–38.

14. Wang J, Knol MJ, Tiulpin A, et al. Gray Matter Age Prediction as a Biomarker for Risk of Dementia. Proc Natl Acad Sci U S A. 2019 Oct 15;116(42):21213–8.

15. Mwangi B, Hasan KM, Soares JC. Prediction of individual subject’s age across the human lifespan using diffusion tensor imaging: a machine learning approach. Neuroimage. 2013 Jul 15;75:58–67.

16. Cole JH. Multimodality neuroimaging brain-age in UK biobank: relationship to biomedical, lifestyle, and cognitive factors. Neurobiol Aging. 2020 Aug;92:34–42.

17. Collins R. What makes UK Biobank special? Lancet. 2012 Mar 31;379(9822):1173–4.

18. Andersson JLR, Sotiropoulos SN. An integrated approach to correction for off-resonance effects and subject movement in diffusion MR imaging. Neuroimage. 2016 Jan 15;125:1063–78.

19. Andersson J, Jenkinson, M., and Smith, S. Non-linear registration aka spatial normalisation. Internal Technical Report TR07JA2, Oxford University, Oxford, UK. 2007a.

20. Peng H, Gong W, Beckmann CF, Vedaldi A, Smith SM. Accurate brain age prediction with lightweight deep neural networks. Med Image Anal. 2021 Feb;68:101871.

21. Simonyan K, Zisserman A. Very deep convolutional networks for large-scale image recognition. arXiv preprint 14091556. 2014.

22. Long J, Shelhamer E, Darrell T, editors. Fully convolutional networks for semantic segmentation. Proceedings of the IEEE conference on computer vision and pattern recognition; 2015.

23. LeCun Y, Bengio Y, Hinton G. Deep learning. Nature. 2015 2015/05/01;521(7553):436–44.

24. Smith SM, Vidaurre D, Alfaro-Almagro F, Nichols TE, Miller KL. Estimation of brain age delta from brain imaging. Neuroimage. 2019 Oct 15;200:528–39.

25. Gottesman RF, Schneider AL, Zhou Y, et al. Association Between Midlife Vascular Risk Factors and Estimated Brain Amyloid Deposition. JAMA. 2017 Apr 11;317(14):1443–50.

26. Bycroft C, Freeman C, Petkova D, et al. The UK Biobank resource with deep phenotyping and genomic data. Nature. 2018 Oct;562(7726):203–9.

27. Oldmeadow C, Holliday EG, McEvoy M, et al. Concordance between direct and imputed APOE genotypes using 1000 Genomes data. J Alzheimers Dis. 2014;42(2):391–3.

28. Du J, Koch FC, Xia A, et al. Difference in distribution functions: A new diffusion weighted imaging metric for estimating white matter integrity. Neuroimage. 2021 Jul 9;240:118381.

29. Imai K, Keele L, Tingley D. A general approach to causal mediation analysis. Psychol Methods. 2010 Dec;15(4):309–34.

30. Franke K, Gaser C. Ten Years of BrainAGE as a Neuroimaging Biomarker of Brain Aging: What Insights Have We Gained? Frontiers in Neurology. 2019 2019-August-14;10(789).

31. Williams OA, An Y, Beason-Held L, et al. Vascular burden and APOE ε4 are associated with white matter microstructural decline in cognitively normal older adults. Neuroimage. 2019 Mar;188:572–83.

32. Wang R, Fratiglioni L, Laukka EJ, et al. Effects of vascular risk factors and APOE ε4 on white matter integrity and cognitive decline. Neurology. 2015 Mar 17;84(11):1128–35.

33. Littlejohns TJ, Holliday J, Gibson LM, et al. The UK Biobank imaging enhancement of 100,000 participants: rationale, data collection, management and future directions. Nat Commun. 2020 May 26;11(1):2624.

34. Koncz R, Sachdev PS. Are the brain’s vascular and Alzheimer pathologies additive or interactive? Curr Opin Psychiatry. 2018 Mar;31(2):147–52.

35. Akinyemi RO, Mukaetova-Ladinska EB, Attems J, Ihara M, Kalaria RN. Vascular risk factors and neurodegeneration in ageing related dementias: Alzheimer’s disease and vascular dementia. Curr Alzheimer Res. 2013 Jul;10(6):642–53.

36. Buford TW. Hypertension and aging. Ageing Res Rev. 2016 Mar;26:96–111.

37. Hamed SA. Brain injury with diabetes mellitus: evidence, mechanisms and treatment implications. Expert Rev Clin Pharmacol. 2017 Apr;10(4):409–28.

38. Debette S, Seshadri S, Beiser A, et al. Midlife vascular risk factor exposure accelerates structural brain aging and cognitive decline. Neurology. 2011;77(5):461–8.

39. Beck D, de Lange AG, Pedersen ML, et al. Cardiometabolic risk factors associated with brain age and accelerate brain ageing. Hum Brain Mapp. 2021 Oct 9.

40. Serrano-Pozo A, Das S, Hyman BT. APOE and Alzheimer’s disease: advances in genetics, pathophysiology, and therapeutic approaches. The Lancet Neurology. 2021 2021/01/01/;20(1):68–80.

41. Suman Kapur SS, Manav Kapoor, Kiran Bala. ApoE Genotypes: Risk factor for Alzheimer’s Disease. Indian Academy of Clinical Medicine. 2006;7(2):118–22.

42. Mirza SS, Saeed U, Knight J, et al. APOE ε4, white matter hyperintensities, and cognition in Alzheimer and Lewy body dementia. Neurology. 2019 Nov 5;93(19):e1807–e19.

43. Schilling S, DeStefano AL, Sachdev PS, et al. APOE genotype and MRI markers of cerebrovascular disease: systematic review and meta-analysis. Neurology. 2013 Jul 16;81(3):292–300.

44. Love S, Miners JS. Cerebrovascular disease in ageing and Alzheimer’s disease. Acta Neuropathol. 2016 May;131(5):645–58.

45. Sweeney MD, Kisler K, Montagne A, Toga AW, Zlokovic BV. The role of brain vasculature in neurodegenerative disorders. Nat Neurosci. 2018;21(10):1318–31.

46. Alqarni A, Jiang J, Crawford JD, et al. Sex differences in risk factors for white matter hyperintensities in non-demented older individuals. Neurobiology of Aging. 2021 2021/02/01/;98:197–204.

47. Palmer BF, Clegg DJ. The sexual dimorphism of obesity. Mol Cell Endocrinol. 2015 Feb 15;402:113–9.

48. Dekkers IA, Jansen PR, Lamb HJ. Obesity, Brain Volume, and White Matter Microstructure at MRI: A Cross-sectional UK Biobank Study. Radiology. 2019 Jun;291(3):763–71.

49. Tankó LB, Bagger YZ, Alexandersen P, Larsen PJ, Christiansen C. Central and peripheral fat mass have contrasting effect on the progression of aortic calcification in postmenopausal women. Eur Heart J. 2003 Aug;24(16):1531–7.

50. Prins ND, van Dijk EJ, den Heijer T, et al. Cerebral small-vessel disease and decline in information processing speed, executive function and memory. Brain. 2005 Sep;128(Pt 9):2034–41.

51. Wallin A, Román GC, Esiri M, et al. Update on Vascular Cognitive Impairment Associated with Subcortical Small-Vessel Disease. J Alzheimers Dis. 2018;62(3):1417–41.

52. Zhang R, Beyer F, Lampe L, et al. White matter microstructural variability mediates the relation between obesity and cognition in healthy adults. Neuroimage. 2018 May 15;172:239–49.

53. Walker KA, Sharrett AR, Wu A, et al. Association of Midlife to Late-Life Blood Pressure Patterns With Incident Dementia. Jama. 2019 Aug 13;322(6):535–45.

54. Marseglia A, Fratiglioni L, Kalpouzos G, Wang R, Bäckman L, Xu W. Prediabetes and diabetes accelerate cognitive decline and predict microvascular lesions: A population-based cohort study. Alzheimers Dement. 2019 Jan;15(1):25–33.

