## Supplementary for "White matter brain age as a biomarker of cerebrovascular burden in the ageing brain"

Supplementary Checklist

### **Supplementary Methods**

Model 1a: WMBAG ~ 𝛽_1_VRS_1 + 𝛽_2_VRS_2 + 𝛽_3_VRS_3 + 𝛽_4_Chronological_age + 𝛽_5_ Sex + 𝛽_6_Scanner + 𝛽_7_APOE + ε_1_

Model 1b: WMBAG ~ 𝛽_1_VRS_1 + 𝛽_2_VRS_2 + 𝛽_3_VRS_3 + 𝛽_4_VRS_1×Sex + 𝛽_5_ VRS_2×Sex + 𝛽_6_VRS_3×Sex + 𝛽_7_Chronological_age + 𝛽_8_ Sex + 𝛽_9_ Scanner + 𝛽_10_APOE + ε_1_

Model 2a: WMBAG ~ 𝛽_1_Hypertension + 𝛽_2_Diabetes + 𝛽_3_Hypercholesterolemia + 𝛽_4_Obesity + 𝛽_5_Smoking + 𝛽_6_ Chronological_age + 𝛽_7_Sex + 𝛽_8_Scanner + 𝛽_9_APOE + ε_1_

Model 2b: WMBAG ~ 𝛽_1_Hypertension + 𝛽_2_Hypertension×Sex + 𝛽_3_ Chronological_age + 𝛽_4_Sex + 𝛽_5_Scanner + 𝛽_6_APOE + ε_1_

Model 2c: WMBAG ~ 𝛽_1_Diabetes + 𝛽_2_ Diabetes ×Sex + 𝛽_3_ Chronological_age + 𝛽_4_Sex + 𝛽_5_Scanner + 𝛽_6_APOE + ε_1_

Model 2d: WMBAG ~ 𝛽_1_Hypercholesterolemia + 𝛽_2_ Hypercholesterolemia ×Sex + 𝛽_3_ Chronological_age + 𝛽_4_Sex + 𝛽_5_Scanner + 𝛽_6_APOE + ε_1_

Model 2e: WMBAG ~ 𝛽_1_Obesity + 𝛽_2_Obesity×Sex + 𝛽_3_ Chronological_age + 𝛽_4_Sex + 𝛽_5_Scanner + 𝛽_6_APOE + ε_1_

Model 2f: WMBAG ~ 𝛽_1_Smoking + 𝛽_2_Smoking×Sex + 𝛽_3_ Chronological_age + 𝛽_4_Sex + 𝛽_5_Scanner + 𝛽_6_APOE + ε_1_

### **Supplementary Tables**

#### **Table e-1 UK Biobank exclusive disease codes**

| Disease | Code |
| --- | --- |
| Stroke or ischaemic stroke | 1081 |
| Transient ischaemic attack | 1082 |
| Subdural haematoma | 1083 |
| Subarachnoid haemorrhage | 1086 |
| Neurological injury/trauma | 1240 |
| psychological/psychiatric problem | 1243 |
| Infections of the nervous system | 1244 |
| Brain/intracranial abscess | 1245 |
| Encephalitis | 1246 |
| Meningitis | 1247 |
| Guillain-Barré syndrome | 1256 |
| Chronic degenerative neurological | 1258 |
| Motor Neuron Disease | 1259 |
| Multiple Sclerosis | 1261 |
| Parkinson’s disease | 1262 |
| Dementia or Alzheimer’s disease | 1263 |
| Epilepsy | 1264 |
| Head injury | 1266 |
| depression | 1286 |
| schizophrenia | 1289 |
| mania/bipolar disorder/manic depression | 1291 |
| Other demyelinating disease | 1397 |
| Cerebral aneurysm | 1425 |
| Cerebral palsy | 1433 |
| Brain haemorrhage | 1491 |
| Spina bifida | 1524 |
| Ischaemic stroke | 1583 |
| Meningioma (benign) | 1659 |

For the reference of cognition standardisation, the participants in UK Biobank with any diseases listed above were removed.

#### **Table e-2 Comparison of characteristics between baseline participants with single time-point MRI scans and participants with repeat scans**

|  | Baseline participants with single time-point scans (n =9759) | Baseline participants with repeat scans (n = 1409) | p value |
| --- | --- | --- | --- |
| ***Demographics*** |  |  |  |
| Chronological age, years, mean ± SD | 64.07 ± 7.56 | 63.05 ± 7.17 | < 0.001 |
| Male, number (%) | 4426 (45.4) | 685 (48.6) | 0.022 |
| Education, college number (%) | 4750 (49.1) | 684 (48.9) | 0.864 |
| ***Vascular risk factors*** |  |  |  |
| HTN, number (%) | 4915 (50.5) | 703 (49.9) | 0.711 |
| Diabetes, number (%) | 544 (5.6) | 66 (4.7) | 0.190 |
| Hypercholesterolemia, number (%) | 2414 (25.0) | 299 (21.4) | 0.003 |
| Obesity, number (%) | 1838 (19.4) | 232 (16.6) | 0.014 |
| Smoking, number (%) | 3763 (38.9) | 459 (32.8) | < 0.001 |
| ***White matter brain age and gap*** |  |  |  |
| White matter brain age, years, mean ± SD | 64.34 ± 8.39 | 62.94 ± 8.02 | < 0.001 |
| WMBAG, years, mean ± SD | 0.28 ± 3.62 | -0.11 ± 3.45 | 0.001 |
| ***Cognition*** |  |  |  |
| Processing speed, z-score, mean ± SD | 0.04 ± 0.98 | 0.28 ± 0.92 | < 0.001 |
| Executive function, z-score, mean ± SD | 0.03 ± 0.99 | 0.24 ± 0.96 | < 0.001 |
| Memory, z-score, mean ± SD | 0.02 ± 1.01 | 0.16 ± 0.97 | 0.015 |
| Global cognition, z-score, mean ± SD | 0.04 ± 0.99 | 0.28 ± 0.93 | < 0.001 |

Chronological age was compared using independent t test, χ^2^ analysis was applied for comparisons of sex, education, and vascular risk factors; ANCOVA analysis was performed for white matter brain age and WMBAG and cognition after controlling for chronological age, sex, scanner and APOE. Abbreviations: ANCOVA = Analysis of Covariance; SD = standard deviation; HTN = hypertension; WMBAG = white matter brain age gap.

#### **Table e-3 Associations between different vascular risk factors and WMBAG change**

|  |  | Unstandardised beta | 95%CI | | p-value |
| --- | --- | --- | --- | --- | --- |
|  |  |  | Lower bound | Upper bound |  |
| **Main effects** | Baseline chronological age | 0.009 | -0.005 | 0.023 | 0.217 |
|  | Sex | -0.204 | -0.401 | -0.006 | 0.043 |
|  | Scanner | -0.285 | -0.391 | -0.179 | < 0.001 |
|  | APOE status | 0.089 | -0.099 | 0.278 | 0.353 |
|  | Hypertension | 0.181 | -0.025 | 0.387 | 0.085 |
|  | Diabetes | 0.322 | -0.154 | 0.798 | 0.185 |
|  | Hypercholesterolemia | -0.200 | -0.465 | 0.066 | 0.140 |
|  | Obesity | -0.371 | -0.644 | -0.099 | 0.008 |
|  | Smoking | -0.054 | -0.262 | 0.154 | 0.612 |
| **Interactions** | Hypertension*Sex | 0.291 | -0.098 | 0.680 | 0.143 |
|  | Diabetes*Sex | 0.316 | -0.639 | 1.270 | 0.516 |
|  | Hypercholesterolemia*Sex | 0.085 | -0.408 | 0.578 | 0.736 |
|  | Obesity*Sex | -0.012 | -0.542 | 0.517 | 0.964 |
|  | Smoking*Sex | 0.194 | -0.215 | 0.603 | 0.353 |

Independent main effects of vascular risk factors on WMBAG change were analysed by adding all vascular risk factors into the regression model. Interaction effects were analysed by adding each vascular risk factor and its corresponding interaction term to the model. Abbreviations: WMBAG = white matter brain age gap; APOE = Apolipoprotein E; CI = confidence interval.

#### **Table e-4 Associations between WMBAG change and cognition change**

|  | Processing speed change | | | | Executive change | | | | Memory change | | | | Global cognition change | | | |
| --- | --- | --- | --- | --- | --- | --- | --- | --- | --- | --- | --- | --- | --- | --- | --- | --- |
|  | Unstandardised beta | 95%CI | | p-value | Unstandardised beta | 95%CI | | p-value | Unstandardised beta | 95%CI | | p-value | Unstandardised beta | 95%CI | | p-value |
|  |  | Lower bound | Upper bound |  |  | Lower bound | Upper bound |  |  | Lower bound | Upper bound |  |  | Lower bound | Upper bound |  |
| Baseline chronological age | -0.006 | -0.012 | < 0.001 | 0.057 | -0.003 | -0.008 | 0.003 | 0.346 | -0.005 | -0.014 | 0.003 | 0.231 | -0.006 | -0.011 | -0.001 | 0.020 |
| Sex | 0.068 | -0.016 | 0.152 | 0.114 | 0.010 | -0.07 | 0.090 | 0.810 | -0.005 | -0.127 | 0.116 | 0.933 | 0.030 | -0.043 | 0.104 | 0.417 |
| Scanner | -0.043 | -0.091 | 0.004 | 0.075 | -0.021 | -0.067 | 0.024 | 0.350 | 0.060 | -0.009 | 0.128 | 0.087 | -0.001 | -0.042 | 0.041 | 0.972 |
| College | 0.024 | -0.059 | 0.108 | 0.567 | 0.052 | -0.028 | 0.132 | 0.202 | -0.053 | -0.174 | 0.068 | 0.394 | 0.014 | -0.059 | 0.087 | 0.703 |
| WMBAG | 0.020 | -0.006 | 0.045 | 0.134 | 0.008 | -0.016 | 0.032 | 0.521 | -0.012 | -0.049 | 0.025 | 0.522 | 0.007 | -0.015 | 0.029 | 0.533 |

This table shows the relationship between WMBAG change and cognition change after controlling for baseline chronological age, sex, scanner, and college. Abbreviations: WMBAG = white matter brain age gap; CI = confidence interval.

#### **Table e-5 Direct and indirect associations between baseline vascular risk factors and longitudinal cognition change**

|  | Processing speed change | | |  | Executive change | | |  | Memory change | | |  | Global cognition change | | |  |
| --- | --- | --- | --- | --- | --- | --- | --- | --- | --- | --- | --- | --- | --- | --- | --- | --- |
|  | Unstandardised beta | 95%CI | | p-value | Unstandardised beta | 95%CI | | p-value | Unstandardised beta | 95%CI | | p-value | Unstandardised beta | 95%CI | | p-value |
|  |  | Lower bound | Upper bound |  |  | Lower bound | Upper bound |  |  | Lower bound | Upper bound |  |  | Lower bound | Upper bound |  |
| ***Direct effects*** |  |  |  |  |  |  |  |  |  |  |  |  |  |  |  |  |
| Hypertension | -0.015 | -0.120 | 0.090 | 0.750 | 0.028 | -0.072 | 0.0130 | 0.590 | -0.048 | -0.198 | 0.090 | 0.500 | -0.015 | -0.108 | 0.080 | 0.760 |
| Diabetes | -0.110 | -0.376 | 0.150 | 0.420 | 0.043 | -0.212 | 0.310 | 0.730 | -0.029 | -0.361 | 0.300 | 0.880 | -0.041 | -0.298 | 0.200 | 0.790 |
| Hypercholesterolemia | 0.018 | -0.128 | 0.160 | 0.800 | 0.001 | -0.140 | 0.140 | 0.990 | -0.028 | -0.094 | 0.040 | 0.420 | -0.049 | -0.173 | 0.080 | 0.470 |
| Obesity | -0.091 | -0.045 | 0.220 | 0.180 | -0.001 | -0.127 | 0.130 | 0.970 | -0.039 | -0.241 | 0.170 | 0.700 | 0.021 | -0.093 | 0.130 | 0.730 |
| Smoking | 0.056 | -0.005 | 0.010 | 0.300 | 0.037 | -0.064 | 0.140 | 0.470 | 0.060 | -0.092 | 0.220 | 0.430 | 0.063 | -0.031 | 0.160 | 0.180 |
| ***Indirect effects mediated through WMBAG change*** | | | |  |  |  |  |  |  |  |  |  |  |  |  |  |
| Hypertension | 0.006 | -0.002 | 0.020 | 0.180 | 0.004 | -0.002 | 0.010 | 0.240 | -0.002 | -0.015 | 0.010 | 0.770 | 0.003 | -0.003 | 0.010 | 0.320 |
| Diabetes | 0.014 | -0.005 | 0.040 | 0.190 | 0.010 | -0.007 | 0.030 | 0.260 | -0.004 | -0.037 | 0.030 | 0.780 | 0.009 | -0.008 | 0.030 | 0.330 |
| Hypercholesterolemia | -0.005 | -0.017 | 0.000 | 0.270 | -0.004 | -0.014 | 0.000 | 0.350 | 0.001 | -0.010 | 0.020 | 0.800 | -0.003 | -0.012 | 0.000 | 0.400 |
| Obesity | -0.085 | -0.024 | 0.000 | 0.140 | -0.006 | -0.020 | 0.000 | 0.220 | 0.002 | -0.016 | 0.020 | 0.800 | -0.005 | -0.019 | 0.000 | 0.300 |
| Smoking | 0.001 | -0.005 | 0.010 | 0.760 | 0.001 | -0.005 | 0.010 | 0.790 | -0.0003 | -0.008 | 0.010 | 0.890 | 0.0007 | -0.004 | 0.010 | 0.800 |

This table shows the direct and indirect associations between baseline vascular risk factors and longitudinal cognitive change mediated through WMBAG change. Chronological age, sex, scanner, and education were controlled for all models. Raw p values were reported in this table with bold unstandardised beta unstandardised beta indicating statistical significance after Bonferroni correction. Abbreviations: WMBAG = white matter brain age gap; CI = confidence interval.

### **Supplementary Figures**

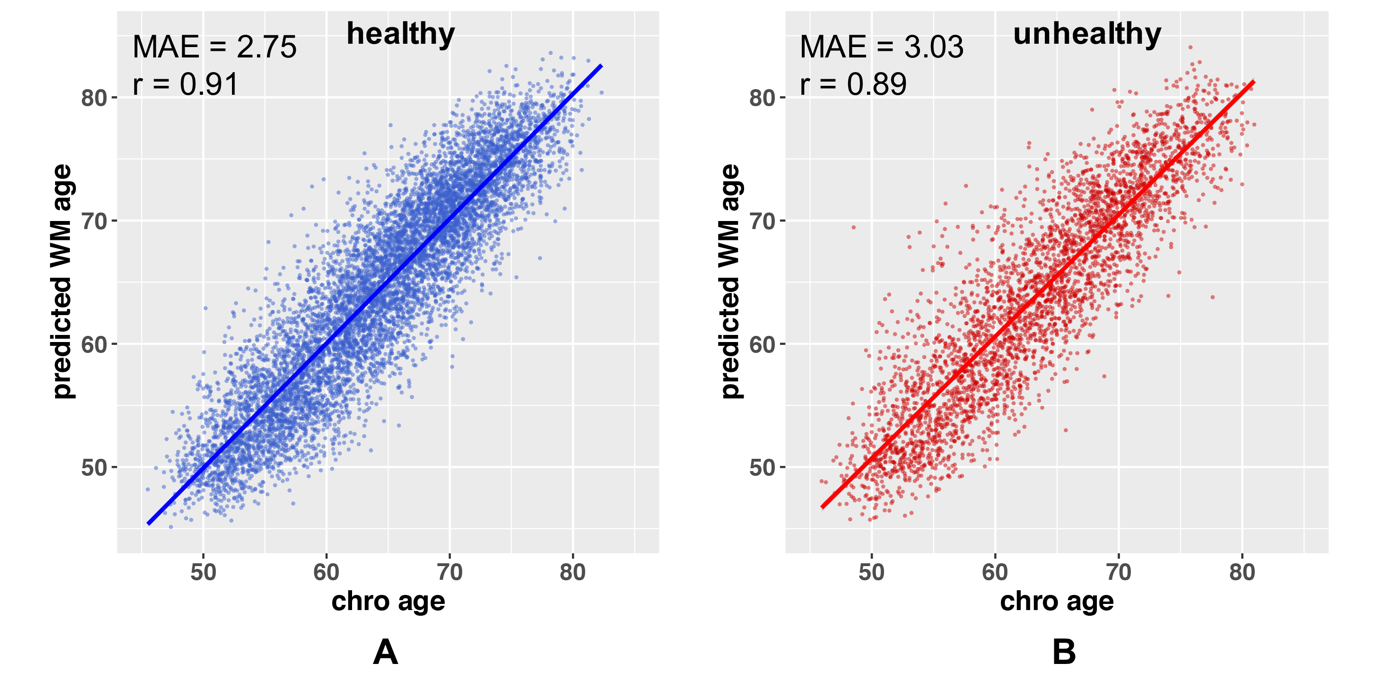

**Figure e-1 Scatterplots showing the relationship between chronological age and predicted white matter brain age in the healthy test set (A) and unhealthy test set (B).** MAE and the correlation coefficient (r) were listed in the upper-left corner of each sub-plot. Abbreviations: r = Pearson’s correlation coefficient; MAE = mean absolute error; WM = white matter; chro age = chronological age.
